# Liver Intrinsic Function Evaluation (LIFE): Multi-parametric Liver Function Profiles of Patients Undergoing Hepatectomy

**DOI:** 10.1101/2024.02.12.24302306

**Authors:** Christian Simonsson, Wolf Claus Bartholomä, Anna Lindhoff Larsson, Markus Karlsson, Shan Cai, Jens Tellman, Bengt Norén, Bergthor Björnsson, Gunnar Cedersund, Nils Dahlström, Per Sandström, Peter Lundberg

## Abstract

**Background & Aims:** For a range of liver malignancies, the only curative treatment option may be hepatectomy, which may have fatal complications. Therefore, an unbiased pre-operative risk assessment is vital, however, at present the assessment is typically based on global liver function only. Magnetic resonance imaging (MRI) modalities have the possibility to aid this assessment, by introducing additional characterization of liver parenchymal, such as non-invasive quantification of steatosis, fibrosis, and uptake function, both for global and regional assessment. To this cause, we here present a prospective observation study (LIFE), in which patients underwent extensive MR-examinations both before and after resective-surgery.

**Approach and Results:** A total of 13 patients undergoing hepatectomy underwent a pre- (n=13) and post (m=5) multimodal MRI examination (within 3-5 days of the surgery) (Fig. 1B). The multimodal MR-examination included DCE, 3D-MRE, fat fraction measurements (PDFF by MRS, 6PD). Using these measurements, we also construct individual patient profiles by including conventional functional, and volumetric measurements, into a multi-parametric space. As a proof of concept, the areas of each profile, denoted ‘multiparametric profile area’ (MPA, and aMPA) were calculated, to create a measurement comprising information from all modalities.

At a group-level, no clear pattern emerged of MPA or aMPA between groups with different extent of resection. In contrast, on a case-by-case basis, several parameters contributed to high individual MPA or aMPA-values, suggesting tissue abnormalities. With respect to regional DCE measurements, *i.e*., relative enhancement at 20 minutes, a clear variation between function in segments, within and between the individuals, was observed.

**Conclusions:** In this combined pre- and post-observational case-based study ranging from very extensive (i) liver surgery to minor (ii), or none (iii), we aimed to describe how a multi-modal MRI examination before hepatectomy could yield valuable information for the pre-operative assessment, with a particular focus on a Couinaud-segmental level. The use of a multi-modal approach allows for a broad spectral characterization of several aspects of the remnant tissue. However, the effectiveness and clinical benefit of each parameter, and how to further optimize an abbreviated clinical MR-protocol needs to be confirmed.

## Introduction

Hepatectomy is in many cases the only treatment for liver malignancies, such as metastasis, *hepatocellular carcinoma* (HCC), or *cholangiocarcinoma* (CCA), and it has the potential to lead to post-hepatectomy liver failure (PHLF). PHLF is a severe complication, with a reported mortality rate of 2% for patients with colorectal metastases and 10% for patients with biliary tumors and HCC [1]. The reported incidence of PHLF is 8-12 %, with higher prevalence in cases with extensive resection [1]. The risk is increased for patients with underlaying chronic liver disease, such as *cirrhosis*, due to impaired remnant functional reserve [2]. Hence, the risk of PHLF is heavily dependent both on the pre-operative liver status as well as the type of malignancy. To minimize the risk of PHLF, a pre-operative assessment of *future liver remnant* (FLR) and liver function is important. To make this assessment a variety of techniques have been deployed [3]. Imaging techniques have shown promise, and a variety of cross-sectional imaging techniques can be used in the context of surgical planning and risk assessment before a hepatectomy [4, 5].

One central imaging technique is magnetic resonance imaging (MRI). Providing high resolution images, MRI has become a clinical standard for evaluation of focal, diffuse, and biliary liver diseases, and it can provide valuable information of liver parenchyma [5]. The spatial resolution provided by MRI (and imaging in general), allows for a non-invasive regional assessment of liver function based on the Couinaud segments. The use of different MRI modalities has shown promise in providing useful information in aiding pre-operative assessment before hepatectomy [6]. There are several MRI modalities that have been studied in the context of pre-operative assessment before hepatectomy.

In the context of pre-operative assessment, one of the most prominent MRI modalities for assessing hepatic function is *dynamic contrast enhanced* (DCE) MRI. For assessing liver function, the hepatocyte specific contrast agent *gadoxetic acid* (Gd-EOB-DTPA, Primovist, Beyer Schering Pharma, Berlin, Germany) is often used. Measures generated from Gd-EOB-DTPA images are based on signal intensities (SI). Some common parameters are e.g., *relative-enhancement* (RE), *liver-to-spleen* (LSC), *liver-to-muscle* (LMC), *hepatocellular uptake index* (HUI), as well as pharmacokinetic-modelling-based parameters [7, 8]. A recent review by Wang *et al*. [9], shows that Gd-EOB-DTPA based parameters have a predictive capacity for PHLF, showing a predictive sensitivity from 75 to 100% and from 54 to 93% in ten reported studies. Moreover, several studies have shown how Gd-EOB-DTPA derived parameters can aid assessment of pre-operative assessment and predict PHLF [10-15].

Magnetic resonance elastography (MRE) is another MR-modality studied in the context of hepatectomy. MRE measures hepatic tissue biomechanical stiffness, which is a well-studied biomarker for *cirrhosis* and can determine the extent of fibrosis. Underlaying cirrhosis has been shown to lead to increased risk of PHLF and post-operative morbidity [16-19], partially due to its correlation with comorbidity factors such portal-hypertension. Stiffness measurements from elastography modalities (via MRE or Ultrasound, *FibroScan*) have been shown to have predictive capabilities of PLHF [20-24]. A recent review by Liang *et al*. [25], showed that preoperative liver-stiffness measured by MRE was a significant predictor of PHLF in patients undergoing liver resection. There are several different implementations options for MRE. The most common implementation is 2D-MRE. However, the more advanced 3D-MRE implementation could in practice yield more detailed tissue assessment, due to improved accuracy. Stiffness, or resistance to deformation, measured by 3D-MRE has been shown to be a valuable biomarker for tumor recurrence after hepatic resection in HCC patients [26-28]. Future studies will be needed to determine any benefit of 3D-MRE in pre-operative assessment before surgery.

The use of a range MR-modalities for the measure of the number of additional biomarkers is relevant for preoperative assessments. One such is the measure of hepatic fat fraction, or PDFF. The presence of steatosis has been shown to increase the risk of postoperative complications [29], with an increased risk of morbidity with *steatohepatitis* (NASH) after resection [30]. Using either ^1^H-MR spectroscopy (MRS) or quantitative mDIXON-sequences, the level of fat infiltration (*proton density fat fraction*, PDFF) can be quantified in the liver parenchyma.

The fat fraction can indicate the levels metabolic dysregulation in the liver. In a study by Sakai *et al*. [31] significantly higher levels of fat fraction were observed in colorectal metastasis patients with tumor recurrence after initial hepatectomy. Additionally, in a study by d’Assignies *et al*.[32] fat fraction was shown to be an independent risk factor for severe complications after hepatic resection. There are other MR-parameters that can contribute to the assessment *e.g*., R2* for *liver iron content* (LIC) assessment, *T1-weighted* (T1w), and *diffusion weighted* (DWI) images. In summary, the mentioned; DCE-MRI, MRE; and fat fraction are three MRI modalities explored in the context of hepatectomy. All yielding independent biomarkers for assessment and prediction of postoperative outcome. Naturally, an MR-protocol combining several MR-modalities and parameters could be of use for the preoperative evaluation.

Our long-term aim is to create a focused and abbreviated multi-modal MR-protocol to aid preoperative assessment for hepatectomy. Here we present the results of a prospective observation study, ‘Liver Intrinsic Function Evaluation’, or LIFE (see an overview in Fig. 1A). A total of 13 patients undergoing hepatectomy underwent a pre- (n=13) and post (m=5) multi-modal MRI examination (within 3-5 days of the surgery) (Fig. 1B). The multimodal MR-examination included DCE, 3D-MRE, fat fraction measurements (PDFF by MRS, 6PD), as well as other sequences (not presented herein). Moreover, due to the personalized nature of each case, the results are presented as a case-by-case study for each patient, ranging from major to minor, surgery, or no surgery, and the contribution of the individual MR-modalities for the assessment pre- and post-hepatectomy.

**Figure 1:**
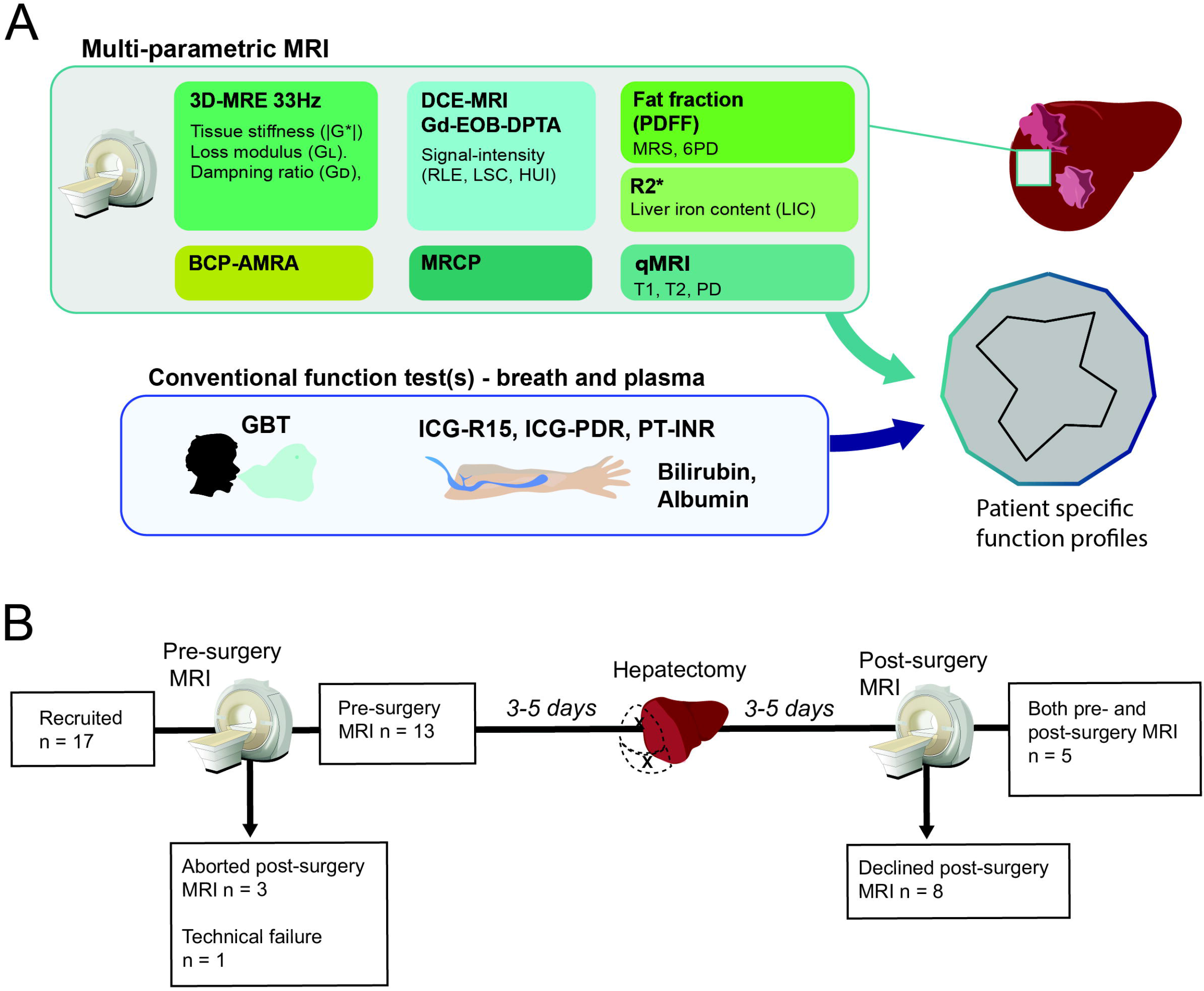
Study-design and the multi-modal magnetic resonance imaging (MRI) examination. **(A)** Multi-modal MR-protocol and conventional function test. The examination can be divided into five segments. The first segment (dark green box) was the elastography (MRE) at 33 Hz vibration frequency, measuring tissue stiffness. The second segment (teal) was dynamic contrast enhanced (DCE) MRI using the hepatocyte specific contrast agent gadoxetic acid (Gd-EOB-DTPA, Primovist, Bayer Schering Pharma, Berlin, Germany). The third box (light green) was quantitative MRI. Fat fraction via both spectroscopy (MRS) and 6-Point Dixon imaging (6PD). Also, the liver iron content (LIC) was estimated using the relaxation rate R2* obtained via standard T2-weighted imaging. We also performed laboratory function tests including galactose breath test (GBT), Indocyanine Green retention rate (R15) and disappearance rate (PDR), as well as measured bilirubin and albumin. **(B)** Study setup. A total of 13 patients, all undergoing hepatectomy, underwent a multi-modal MRI examination 3-5 days before and after surgery. A total of five patients were able and willing to undergo the post-surgery MRI examination.

## Materials and Methods

### Study Setup and Cohort

A total of 17 patients scheduled for partial hepatectomy were recruited on referral from the Linköping University Hospital, Linköping, Sweden between the years 2014 – 2017. The cohort characteristics can be found in Table 2. Four patients obtained postoperative chemotherapy, one recurrence, and the 90 days mortality in the cohort was two patients (see details in Table 1).

**Table 1:**
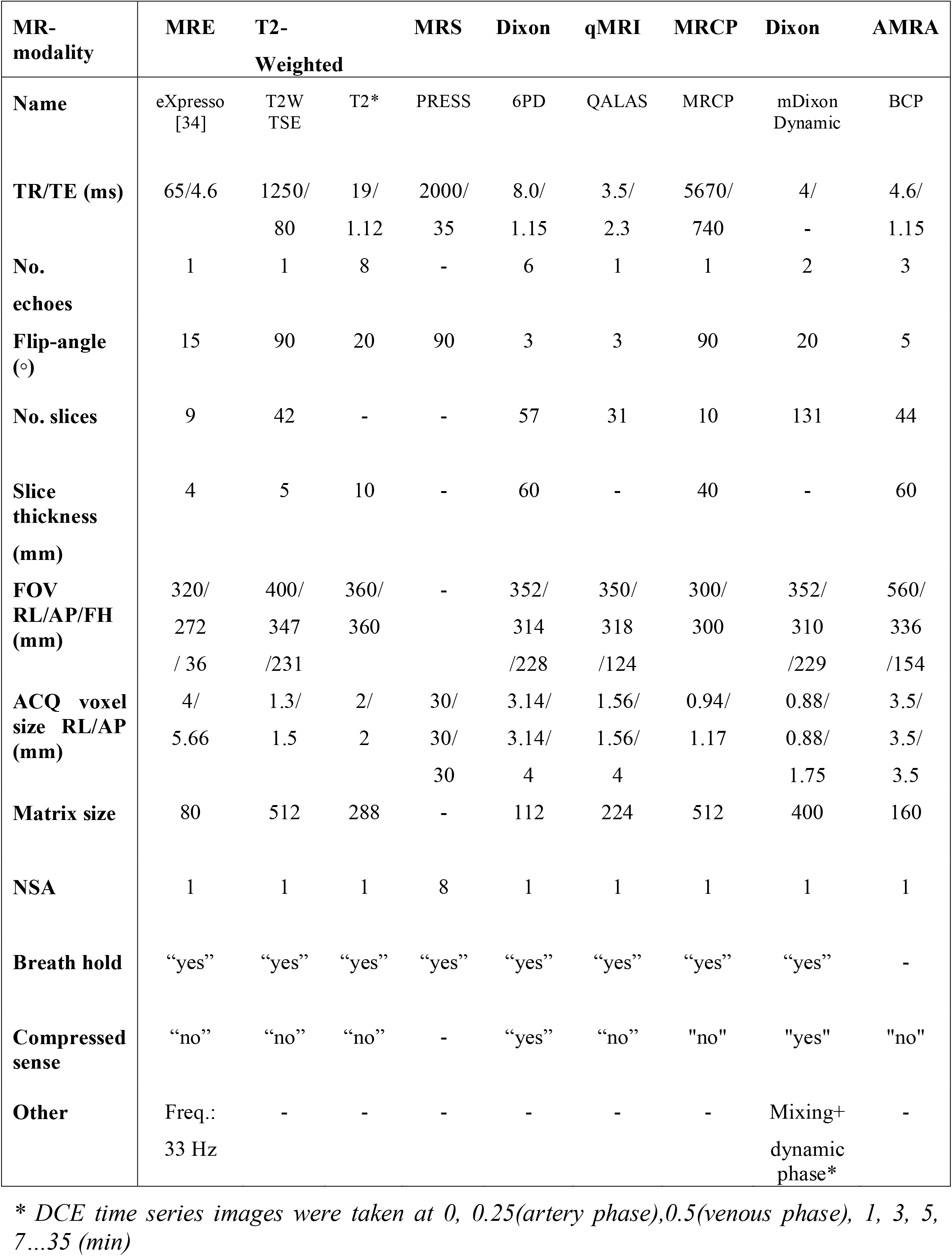
The LIFE multi-modal MR-protocol. This provides an overview of the sequences and sequences parameters included in the MR-protocol. The sequences are presented in sequential order.

**Table 2:**
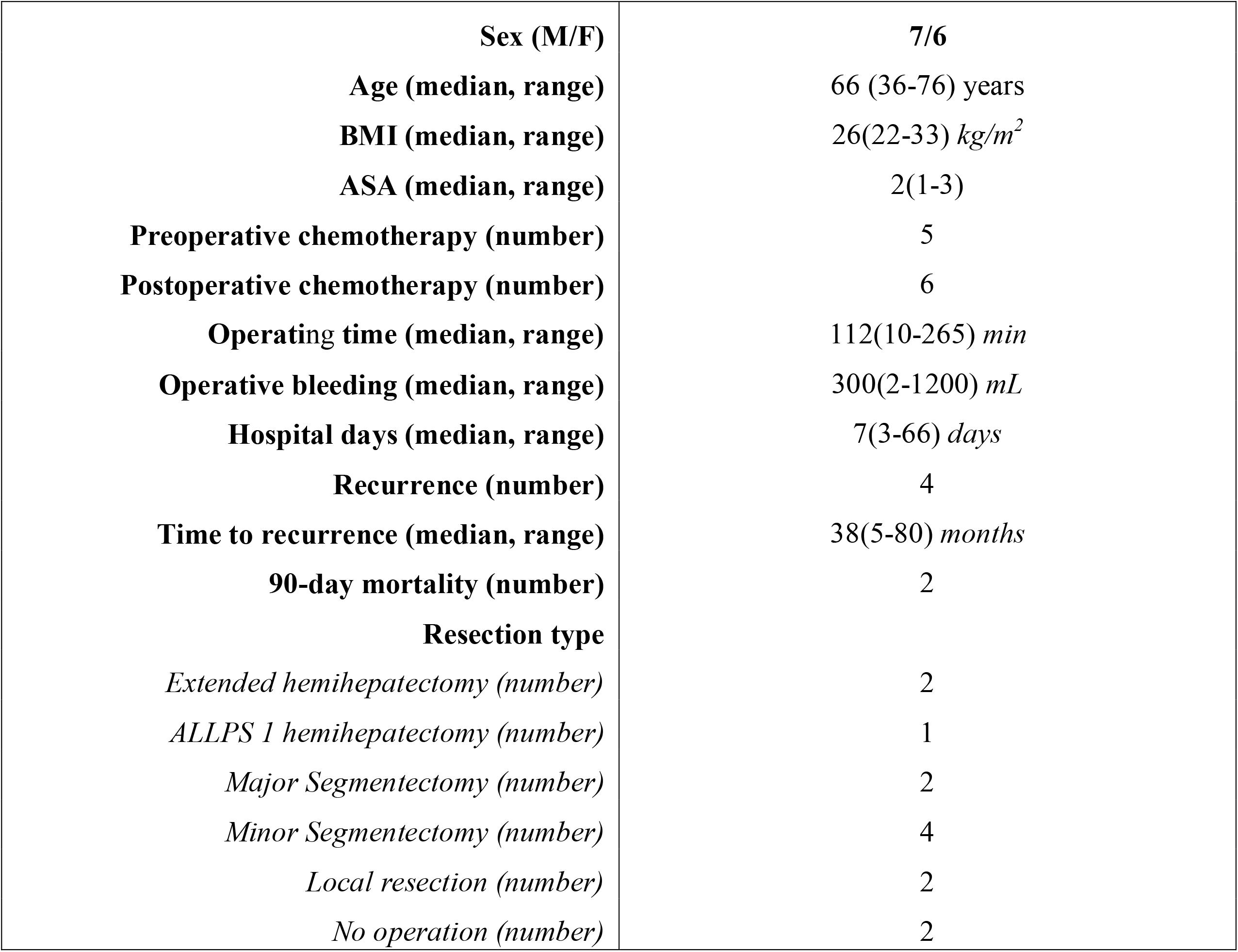
The LIFE cohort. Table describing general cohort characteristics and surgical parameters.

The study pipeline included two separate and subsequent MR-examinations; one pre- and one post-surgery. Three patients were excluded due to aborting the pre-surgery MRI examination, and one patient was excluded due to technical failure. Pre-surgery data was therefore collected for a total of 13 patients. Nine patients declined the follow-up post-surgery MR-examination for personal reasons. Thus, a total number of five patients underwent both pre-and-post-surgery MRI (Fig. 2A-B). All patients also underwent the clinical standard assessment of surgical feasibility, and functional hepatic reserve. The local ethics committee approved this study, and written informed consent was obtained from all patients (Dnr. M160-09 and rev100615). Patients are numbered and grouped based on the type of resection in the manuscript.

**Figure 2:**
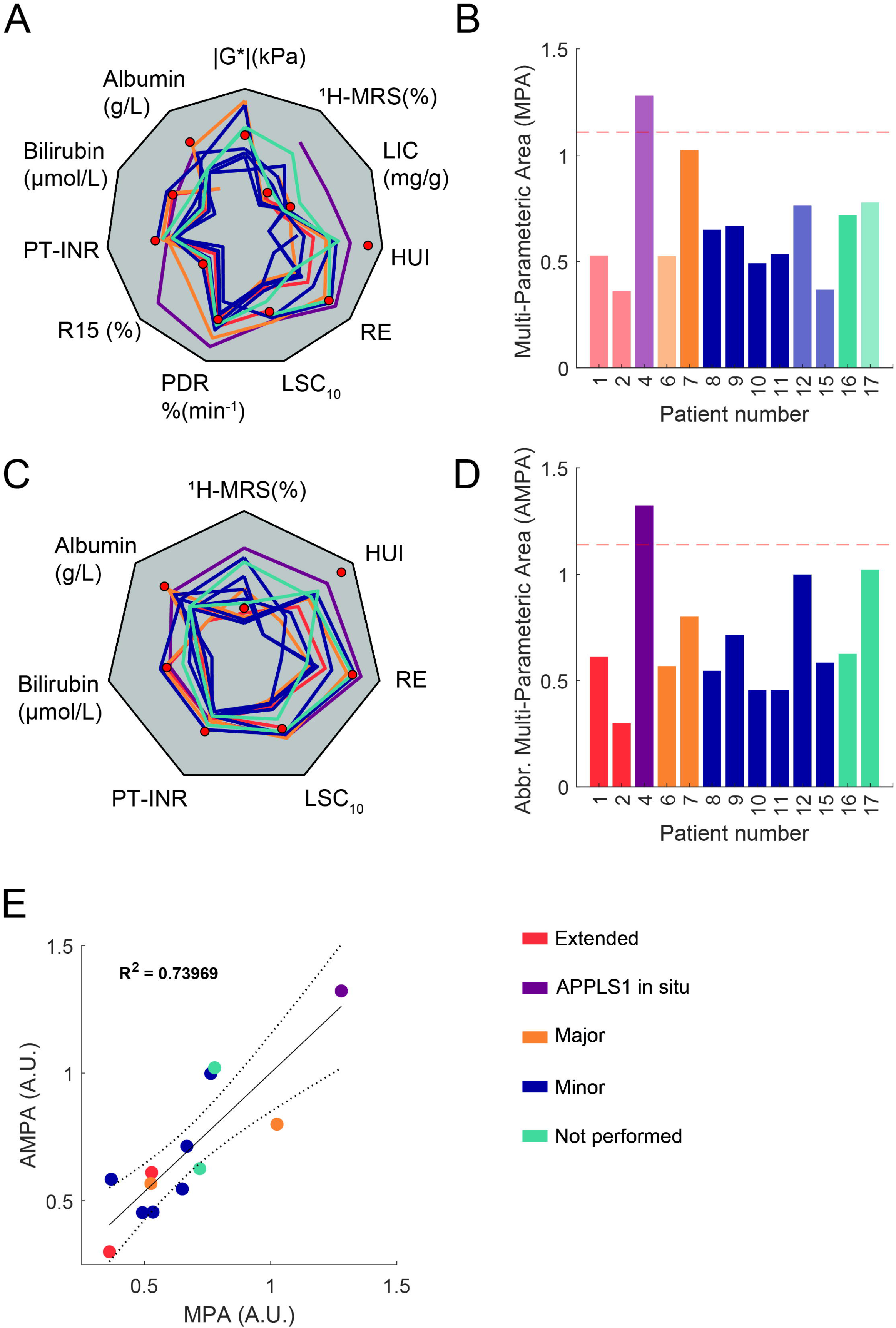
Multi-parametric profiles of pre-surgery MRI examination analysis. **(A)** Radar chart over all MR-modalities and the resulting multi-parametric space before surgery(pre-op). Each line represents the patient multiparametric profile using different MRI parameters as dimensions. The parameter profile which also included additional plasma-based measures; function test using indocyanine green (ICG) retention rate after 15 min (R15) and plasma disappearance rate (PDR), as well plasma concentration of bilirubin and albumin, and lastly prothrombin time international normalized ratio (PT-INR). The red dots are literature-based threshold values for all different measures. The line is broken when the data is unavailable for the specific patient. The color of the line indicates the extent of resection the patient went through; extended (red), ‘Associating Liver Partition and Portal vein Ligation for Staged’ hepatectomy ALPPS1 (purple), major (orange), minor (blue), and surgery not performed (dark teal). The red dots are reference values indicating e.g., presence of steatosis, iron overload, fibrosis, and reduced liver uptake function. **(B)** Bar plot showing the values of the multi parametric area (MPA) for all patients. Patients are numbered along the x-axis. The faded bars i.e., patient1, indicates that values that not all parameters were available. The red line indicated the threshold value calculated based on reference values. The area created by each parameter profile can be used as an indicator of total liver status. The axis was flipped for measures, where low values indicate lowered liver function. The axis where also normalized so that all measures held the same weight when calculating the area measurement. The weight for each measure can be adjusted. The red dotted line indicates the area created by the literature-based thresholds. The faded bars represent patients where not all data was available. **(C)** Here the abbreviated multi-parametric profiles are shown, where only measures which were available in all patients were included. The main difference is the removal of MRE, liver iron content, and the ICG based plasma measurements. **(D)** Bar plot showing abbreviated multi-parametric profile (aMPA) areas values. **(E)** Regression plot between the MPA and aMPA. The line is fitted liner regression, and 95% confidence bounds, yielding the correlation R^2^= 0.74.

### 3D Magnetic Resonance Elastography

The 3D-MRE method has been described previously in [33]. The 3D-MRE was done via the eXpresso [34] sequences and the used sequence parameters can be found in Table 4. An active electrodynamic transducer (Philips Medical, Hamburg, Germany) was used to transmit mechanical waves at 33 Hz on the right side of the patient at the level of the liver, with the patient in the supine position. Images were then acquired using four separate breath-holds, and nine slices were acquired with a slice thickness of 4 mm each. Image post-processing was done using in-house software. Regions of interests (ROIs) drawn by an experienced operator (CS) with support from an experienced radiologist (ND), following the QIBA guidelines. The ROIs were drawn using the magnitude image, and shear-wave quality was checked using the phase-wave image, as well as the signal-to-noise ratio (SNR), and divergence from sine wave was checked.

### ^1^H-MRS and 6-point-Dixon)

Liver PDFF was measured using ^1^H-MRS, specifically a PRESS sequence as described previous publications [35]. The sequences parameters can be found in Table 1. The spectral voxel placement was done by an experienced radiographer. Post-processing of the MRS data was performed by quantifying the integrals of water and fat resonances, respectively, using jMRUI and the AMARES algorithm [36, 37]. The integrals were also corrected for differences in T_1_ and T_2_. The T_1_ relaxation times for fat and water were assumed to be 236 and 663 ms, respectively [38]. The T_2_ relaxation time for was assumed to be 58.5 ms [39], and the T_2_ relaxation time for water was assumed to be 43.4 ms [40].

Liver MRI-PDFF was measured was also measured using a six-point Dixon (6PD) sequences. The sequence parameters can be found in Table 1. The ROI placement was done by an experienced radiologist (WB).

### Quantitative MRI – qMRI

We also used Quantitative MRI (qMRI) for rapid determination of R1, R2 and proton density (PD) in a single breath-hold using the 3D-QALAS sequence[41]. The sequence parameters are shown in Table 1.

### Whole Body MRI (BCP)

We also collected whole-body composition imaging using Dixon-sequences using the AMRA protocol, images where automatically segmented using the AMRA Research (AMRA Medical AB, Linköping, Sweden) postprocessing software.

### T2*, R2* and calculations of liver iron content (LIC)

The method of acquisition and quantification of T2* and R2* has previously been described in [42]. The acquisition was done via an axial 3D multi-echo field echo sequence with spectral pre-saturation with inversion recovery fat saturation. Sequences parameters can be found in Table 1. For the quantification of R2* a ROI was placed in the area in the liver of interest by an experienced radiologist (WB), blinded to all other study results. The value of R2* was calculated by fitting a mono-exponential decay function to the mean signal intensities from the ROIs at the different echo times. The liver iron content (LIC) was calculated according to Wood’s formula implemented for 3 T [43]. The formula used (for 3 T) was defined in the following way:

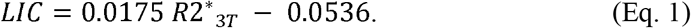

### Dynamic contrast enhanced MRI

Magnetic Resonance was performed using a 3.0 T Philips Ingenia MR-scanner (Philips Healthcare, Best, The Netherlands) and a phased-array body coil. Following a bolus injection of Gd-EOB-DTPA (0.025 mmol/kg body weight), images were acquired using an axial breath-hold, fat-saturated, T1-weighted, 3D gradient echo sequence. Typical image parameters included flip angle: 10°, repetition time: 4.2 ms, echo time: 2.0 ms, SENSE factor: 1.7, field of view: 300×200×350 mm^3^. The post□injection images included arterial and portal venous phases, as well as time-series images acquired *e.g*., between 0, and 50 minutes. An example of acquired images are shown in Fig. 2C.

In post-processing ROIs were placed in each of the eight *Couinaud* segments, plus three ROIs in the spleen, as well as ROIs in muscle, and portal vein by an experienced radiologist (BN and WB). Signal intensity measurements were obtained from each ROI. From the signal intensity the T1 relaxation rate (R1).

### Calculations of Signal Intensity Contrast Measurements

We used several different phenomenological calculations of liver function based on the signal intensity as described in [44]. These calculations were performed both for estimating global liver function, but also for each *Couinaud* segment. This section will go through each calculation. The hepatocellular uptake index (HUI) at 20 minutes after contrast injection defined as

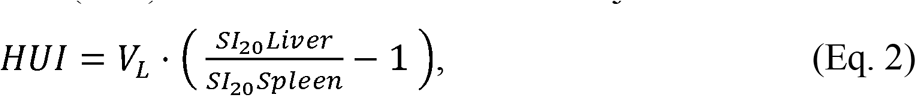

where *V*_*L*_ is the total liver or segment volume, *SI*_20_ is the signal intensity at 20 minutes for either liver or spleen. The relative enhancement (RE) of the liver was calculated as

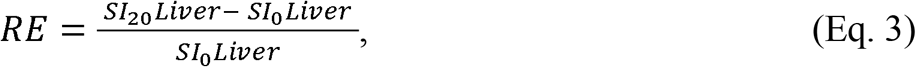

where *SI*_0_ is the signal intensity at time point 0. The liver-to-spleen contrast ratio (LSC) was calculated as

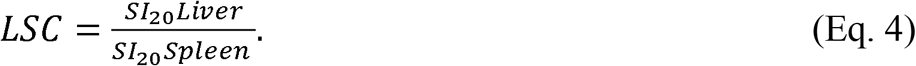

The LSC was also calculated at 10 minutes, as was the normalized LSC which can be written as

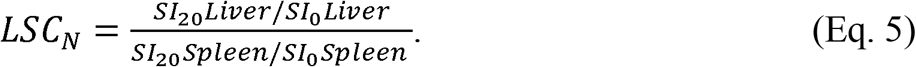

In addition, the liver-to-muscle contrast ratio was also calculated, written as

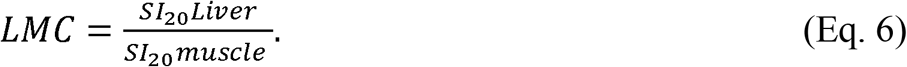

Finally, we also calculated the area under curve (AUC) for each of the liver SI time-series curves.

### Calculation of multi-parametric area profiles

The multi-parametric area profiles (MPA) were determined by measuring the area of each patient profile in the multi-parametric space. The multi-parametric space has several axes representing all individual parameters, organized after category. The axes limits were normalized, in such a way that all parameters were equally scaled (axes were set to origo, equal to 0 and the max value being 1). For parameters where a lower value indicates tissue dysfunction the axis was inverted. In a few cases where certain parameter values were absent, the parameter values were set to zero, so as not to contribute to the MPA. In contrast, in the abbreviated MPA (aMPA), we only included values that were present for all patients.

The values used to calculate the suggested cut-off were the following: ^1^H-MRS >3 % PDFF [35] for presence of steatosis, *liver iron content* (LIC) > 1.8 [45, 46] for low liver iron overload, Hepatocellular uptake index (HUI) 573.44 [47] for indication of PHLF(98% sensitivity; 83% specificity), *relative enhancement* (RE) <0.9 [48] cut-off for presence of fibrosis, liver-to-spleen contrast ratio at 10 minutes <1.24 [49] cut-off for presence of fibrosis, R15 > 10 % and PDR < 19 %·min^-1^ [50, 51], PT-INR, bilirubin, and albumin where all based on the Child-Pugh classification, and lastly no literature threshold exist for 3D-MRE stiffness (neither at 33 Hz, or any other frequencies). These cut-offs are indicated as red dots in Fig. 2B.

### Conventional Function Tests

In connection to each MR-examination a laboratory examination was also done, where plasma samples were collected, and conventional functional lab tests were performed.

#### Galactose Breath Test (GBT)

After an overnight fast, the patients drank a solution consisting of 0.5 g/kg body weight unlabeled galactose together with 5 mg/kg body weight [1-^13^C]-labeled galactose dissolved in water (2 mL/kg body weight). Breath tests were collected with the patient at rest before and 30, 60, 90, 120, 150 and 180 minutes after drinking the spiked solution. Breath samples were analyzed for ^13^CO□ by isotope ratio mass spectrometry, reference value 10.8% – 18.6%. The results were presented as the cumulative percentage of the administered dose of ^13^C-galactose.

#### Indocyanine Green-test (ICG)

A bolus dose of 0.5 mg/kg body weight indocyanine green (ICG) was injected into a peripheral vein. Through a finger piece sensor, the ICG retention rate (ICG-R15) and ICG plasma disappearance rate (ICG-PDR) were evaluated with the non-invasive pulse-densitometric LIMON system (Impulse Medical System, Munich, Germany).

#### Blood Samples and Weight

Each test was performed pre- and post-operatively either the day before or the day after MRI to eliminate any confounding effects on the MRI measurements, blood samples (PT-INR, Bilirubin and Albumin) and body weight were also collected in connection with these examinations.

## Results

### Multi-Modal MR-Protocol Results in a Multi-Parametric Space for Pre-Operative Assessment

The analysis of the MR-data gathered before surgery (pre-op) resulted in a wide range of independent parameters for global assessment of liver status. The multi-parametric space is constituted of parameters covering three different modalities: DCE-MRI, 3D-MRE and quantitative MRI. The individual patient profiles, indicating the extent of resection each patient underwent; extended, ALPPS1(*in situ* split), major, minor, or not performed. The multi-parametric space thus summarizes the information of several important aspects of the status of liver parenchyma; fat fraction and the level of steatosis, 3D-MRE stiffness which correlates to degree of fibrosis and contrast uptake ability.

All information from the multi-parametric space was additionally also compressed into a singular comprehensive view, which we denoted ‘*abbreviated parameter profiles*’. These profiles are shown as radar charts. A sub-set of parameters, at least one from each separate MRI-modality, were then selected and combined with established conventional plasma-based measurements of liver function. The included measurements were the following: function test using *indocyanine green* (ICG) retention rate after 15 min (R15, %) and plasma disappearance rate (PDR, %·min^-1^), as well plasma concentration of bilirubin (μmol·L^-1^) and albumin (g·L^-1^), and lastly *prothrombin time international normalized ratio* (PT-INR). For comparison, literature cut-off values for each measurement were included, and the cut-off values were detailed above.

Using each profile, single scalar values were thus constructed, based solely on the appearance of the individual profiles, and they were thus determined for each patient. As described above, the chart axis was also normalized (or reversed as required) in such a way that an increase in area was indicative of reduced liver functionality, all measurement contributes equally to the area. The areas were denoted as the ‘*multi-parametric area*’ (MPA), and they were determined for each individual patient separately. The literature-based threshold values (MPA = 1.11) were calculated and added to the graphs to simplify comparison. Only patient 4 was above the threshold with MPA = 1.28. An abbreviated version of MPA profiles was also defined and denoted aMPA. Interestingly, the profiles follow a similar order of magnitude, with patient 4 still having the highest value.

In the section below, each patient is discussed individually with respect to their (a)MPA-values, pre-operative assessment, starting with patients for which two or more segments were resected, and these were classified as belonging to the *‘major resection’* category.

### Multi-Parametric Profiles for Patients Undergoing Major Resections

A total of seven patients were in the major resection category: Patients 1, 2 (*extended*), 4 (*ALPPS*), 6 and 7 (*major*). Patient MRI data from pre-and post-surgery can be found in Table 3. In the following section each patient is described separately.

**Table 3:** MRI based measures for the major resection group.

| **Patient** | **1** | **2** | **4** | **6** | **7** |
| --- | --- | --- | --- | --- | --- |
| **Cancer diagnosis** | β-cell lymphoma | Bile duct | Colonic metastasis | Colorectal metastasis | Colonic metastasis |
| **Resection** | 4B, 5, 6, 7, 8. | Half of 1, majority of 4A, 4B, 5, 6, 7 and 8. | 4B and 5 (p1)<br>6, 7, and 8. (p2) ** | Majority of 4, part of 6, 2 and 3 | 2, 3, 4A |
| **Complication** | PHLF | † pod 8<br>MOF | Suspected<br>PC | N/A | N/A |
| **<sup>1</sup>H-MRS (PDFF) (%)** | 1.2 | 0.9 | 6.8 | 3.3 | 0.8 |
| **Pre-op 6PD (PDFF) (%)** | 0.2 | -4.8 | 13.1 | 6.1 | 1.6 |
| **Post-op 6PD (PDFF) (%)** | 3.7 | N/A | N/A | 8.4 | -0.7 |
| **Pre-op R2\*(s<sup>-1</sup>)** | 35.8 | 60.1 | 167.3 | 70.5 | 44.0 |
| **Post-op R2\*(s<sup>-1</sup>)** | 42.8 | N/A | N/A | 46.6 | 51.0 |
| **Pre-op LSC<sub>10</sub>** | 3.0 | 1.6 | 1.8 | 3.3 | 2.1 |
| **Post-op LSC<sub>10</sub>** | 1.3 | N/A | N/A | 1.8 | 1.7 |
| **Pre-op LSC<sub>20</sub>** | 3.5 | N/A | 2.0 | 3.8 | 2.2 |
| **Post-op LSC<sub>20</sub>** | 1.4 | N/A | N/A | 1.9 | 1.9 |
| **Pre-op NLSC<sub>10</sub>** | 1.8 | 1.41 | 1.2 | 1.9 | 1.2 |
| **Post-op NLSC<sub>10</sub>** | 1.1 | N/A | N/A | 1.3 | 1.2 |
| **Pre-op NLSC<sub>20</sub>** | 2.1 | N/A | 1.3 | 2.2 | 1.3 |
| **Post-op NLSC<sub>20</sub>** | 1.2 | N/A | N/A | 1.4 | 1.4 |
| **Pre-op LMC<sub>20</sub>** | 3.6 | N/A | 2.5 | 3.8 | 2.1 |
| **Post-op LMC<sub>20</sub>** | 1.6 | N/A | N/A | 1.8 | 1.6 |
| **Pre-op RE<sub>20</sub>** | 1.5 | N/A | 0.8 | 1.7 | 1.1 |
| **Post-op RE<sub>20</sub>** | 0.5 | N/A | N/A | 0.8 | 1.1 |
| **Pre-op HUI<sub>20</sub>** | 3316 | N/A | 1543 | 4393.4 | 2697 |
| **Post-op HUI<sub>20</sub>** | 251 | N/A | 1251 | N/A | 1630 |
| **\|G\*\| (kPa)** | N/A | N/A | N/A | N/A | 1.34 |

\*\* APPLS two-part p1 & p2

**Patient 1** participated both in the pre-and post MRI examination. Patient 1 underwent extended resection and developed PHLF. Both ^1^H-MRS, and 6PD PDFF were low (1.2 and 0.6%) indicating no steatosis. MRE stiffness values were not available, and no significant iron content was found either pre-or post-surgery (0.57 and 0.69 mg/g). None of the SI contrast measurement were low pre-surgery (e.g., RE = 1.5 > 0.9, NLSC_10_ = 1.8 > 1.24). However, after surgery all contrast SI measurements were significantly lowered e.g., HUI = 251.3 (Table 3). A more detailed regional analysis of all resected and remnant segments could not be done, a possible cause was the small remnant liver volume.

**Patient 2** also underwent an extended resection, and later developed MOF and passed away POD 8. The patient had *HCC* severe liver cirrhosis, chronic inflammation. The Gd-EOB-DTPA DCE imaging was aborted before the 20 minute mark due to nausea, thus many MR-contrast agent based measures were unavailable. There was no presence of steatosis (<0% 6PD and 0.9% MRS PDFF), as well as no excess iron content (0.99 mg/g), and slightly lower NLSC_10_ (1.41).

**Patient 4** had both the highest MPA and aMPA value. Patient 4 had suspected PC. In addition, the patient had the highest PDFF (6.8% ^1^H-MRS and 13.1% 6PD) indicative of developed steatosis, and the highest LIC (2.87 mg/g) indicative of mild iron content. The patient also had low SI based measures (HUI = 1251, NLSC_10_ = 1.2, and RE = 0.8). In addition, the patient also had the most extreme ICG-measures (R15 = 19.8 % and PDR = 10.8). Moreover, this patient was also the only with a *Clavien-Dindo score* for surgical complications of 5, as well as the longest hospital stay (66 days) and later passed away POD 56 (*peritonitis* with multiorgan failure). The patient did, for these reasons, not undergo the post-surgery control MR-examination, but ICG functional tests were nevertheless performed (R15 = 17.8% and PDR = 11.5%·min^-1^), showing lower clearance of ICG.

**Patient 6 and Patient 7**. The last two patients both underwent the post-surgery MRI examination. Patient 6 had steatosis (3.3% ^1^H-MRS and 6.1% 6PD), while patient 7 had none (0.8% ^1^H-MRS and 1.6% 6PD). Patient 7 had a slightly higher aMPA value (0.8) compared to patient 6 (0.57). Patient 7 had stiffness assessed by 3D-MRE and had a higher value than the remainder of the cohort (1.34 kPa).

### Multi-Parametric Profiles for Individual Patients Undergoing Minor-to-No Resection

In the minor and no-resection category a total of eight patients were included, where patients 8, 9, 10, 11, 12 and 15 underwent minor resection, and patients 16 and 17 did undergo surgery, but not any resection. MR-data from pre-and post-surgery can be found in Table 4. In the following section each patient is described separately.

**Table 4:** MRI based measures for the minor resection group.

| Patient | 8 | 9 | 10 | 11 | 12 | 15 | 16 | 17 |
| --- | --- | --- | --- | --- | --- | --- | --- | --- |
| **Cancer diagnosis** | Benign | Focal nodular hyperplasia | Colorectal metastasis | Colonic metastasis | Benign | Colonic metastasis | Sarcoma | Bile duct |
| **Resection** | Part of 6 | 2, 3 | 3 | Part of 6 and part of 8 | Part of 6 | 2,3 and part of 7, 6, 5, 4B | - | - |
| **Complication** | - | Fluids at resection edge | - | Gastric retention |  | Pneumonia | - | - |
| **<sup>1</sup>H-MRS (PDFF) (%)** | x | 0.3 | 4.4 | 3.2 | 0.6 | 6.0 | 2.0 | 5.6 |
| **Pre-op 6PD (PDFF) (%)** | 3.2 | -0.3 | 6.6 | 5.5 | -0.0 | 7.1 | 2.7 | 5.0 |
| **Post-op 6PD (PDFF) (%)** | 1.8 | N/A | N/A | N/A | 1.8 | N/A | N/A | N/A |
| **Pre-op R2\*(s<sup>-1</sup>)** | 50.1 | 57.6 | 53.1 | 61.5 | 47.4 |  | 49.7 | 94.5 |
| **Post-op R2\*(s<sup>-1</sup>)** | 34.5 | N/A | N/A | N/A | 64.0 | N/A | N/A | N/A |
| **Pre-op LSC<sub>10</sub>** | 2.86 | 2.29 | 2.77 | 2.76 | 1.84 | 3.11 | 2.85 | 2.13 |
| **Post-op LSC<sub>10</sub>** | 2.39 | N/A | N/A | N/A | 1.08 | N/A | N/A | N/A |
| **Pre-op LSC<sub>20</sub>** | 3.46 | 2.85 | 3.49 | 3.10 | 1.90 | 3.97 | 2.93 | 2.42 |
| **Post-op LSC<sub>20</sub>** | 2.55 | N/A | N/A | N/A | 1.06 | N/A | N/A | N/A |
| **Pre-op NLSC<sub>10</sub>** | 1.75 | 1.28 | 1.86 | 1.87 | 1.27 | 1.85 | 1.57 | 1.33 |
| **Post-op NLSC<sub>10</sub>** | 1.73 | N/A | N/A | N/A | 1.11 | N/A | N/A | N/A |
| **Pre-op NLSC<sub>20</sub>** | 2.13 | 1.59 | 2.34 | 2.11 | 1.32 | 2.36 | 1.61 | 1.51 |
| **Post-op NLSC<sub>20</sub>** | 1.85 | N/A | N/A | N/A | 1.09 | N/A | N/A | N/A |
| **Pre-op LMC<sub>20</sub>** | 3.51 | 3.05 | 3.31 | 2.52 | 1.79 | 3.37 | 2.95 | 2.37 |
| **Post-op LMC<sub>20</sub>** | 2.54 | N/A | N/A | N/A | 1.20 | N/A | N/A | N/A |
| **Pre-op RE<sub>20</sub>** | 1.67 | 1.34 | 1.60 | 1.66 | 0.93 | 2.23 | 1.79 | 0.977 |
| **Post-op RE<sub>20</sub>** | 1.44 | N/A | N/A | N/A | 0.57 | N/A | N/A | N/A |
| **Pre-op HUI<sub>20</sub>** | 3763 | 2382 | 5433 | 5443 | 2220 | 4075 | 2110 | 2546 |
| **Post-op HUI<sub>20</sub>** | 2592 | N/A | N/A | N/A | 85.1 | N/A | N/A | N/A |
| **\|G\*\| (kPa)** | 0.67 | 0.62 | 0.44 | 0.72 | 1.28 | N/A | 0.98 | 1.0 |
| **G<sub>D</sub> (kPa)** | 0.56 | 0.53 | 0.40 | 0.60 | 0.93 | N/A | 0.81 | 0.85 |
| **G<sub>L</sub> (kPa)** | 0.24 | 0.20 | 0.11 | 0.23 | 0.49 | N/A | 0.36 | 0.37 |
| **alpha (1/m)** | 0.055 | 0.055 | 0.057 | 0.055 | 0.058 | N/A | 0.052 | 0.050 |
| **c (s<sup>-1</sup>)** | 0.81 | 0.78 | 0.67 | 0.84 | 1.14 | N/A | 1.00 | 1.01 |

**Patient 8** was on the border for steatosis (3.19% 6PD and 2.38% MRS PDFF). The iron content (0.82 mg/g < 1.8 mg/g) was also low. None of the SI based measured indicated impaired uptake capabilities (see Table 4). Additionally, the 3D-MRE biomechanical parameters did not show the presence of fibrosis. No pre-surgery MRI data showed any pathological values. The ICG-PDR was below the threshold indicating slower clearance of ICG (17.6%·min^-1^). Patient 8 was in the lower span of MPA and aMPA (0.64 and0.54).

**Patient 9** had low both PDFF and LIC. Certain SI measurements were low, e.g., NLSC_10_=1.28. The 3D-MRE biomechanical parameters did not show any signs of the presence of fibrosis. ICG parameters did not indicate any tissue dysfunction (PDR = 29.1%·min^-1^ and R15 = 1.3%). Patient 9 was in the lower span of MPA and aMPA (0.67 and 0.71).

**Patient 10** had slight steatosis (6.57% 6PD and 4.35% MRS PDFF), but no other measure indicated reduced liver function. Patient 10 was in the lower span of MPA and aMPA (0.49 and 0.45).

**Patient 11** also had slight steatosis (5.54% 6PD and 3.21% MRS PDFF), but no other measure indicates reduced liver function. Patient 11 was in the lower span of MPA and aMPA (0.53 and 0.46).

**Patient 12** had a benign tumor, but still had a high MPA and aMPA (0.76 and 0.99). No presence of steatosis was found, and the iron content was also low (LIC = 0.77 mg/g). No change was seen in PDFF or LIC after surgery (see Table 4). However, the patient had extensive cirrhosis which was reflected in the high 3D-MRE stiffness (|G*| =1.28 kPa). As expected, the SI measurements were also low e.g., RE = 0.93, NLSC_10_ = 1.08. And after surgery, the SI measures where even lower, e.g., RE = 0.93, HUI = 85.1.

**Patient 15** had clear signs of steatosis (5.96% 6PD and 7.11% MRS PDFF). However, no other measures indicated any tissue dysfunction.

**Patient 16** did undergo surgery but did not undergo any resection (Table 4). There was no presence of steatosis (2.72 % 6PD and 2.02 % MRS PDFF) and also low LIC (0.82 mg/g). The 3D-MRE stiffness (|G*| =0.98 kPa) was in the high range in the cohort, which might be indicative of the presence of some fibrosis.

**Patient 17** had PC and had surgery but did not undergo resection. The patient had an MPA and aMPA of 0.77 and 0.99 respectively. There was presence of a degree of steatosis (4.97% 6PD and 5.59% MRS PDFF) and LIC was also 1.60 mg/g. The 3D-MRE stiffness was in the upper range in our cohort (|G*| =1.0 kPa). Some of the SI measures were low e.g., RE = 0.98.

### Abbreviated MPA Suggests fibrosis

A total of eight patients underwent 3D-MRE during the pre-surgery MRI examination (Table 3-4). From previous studies [52], we know that SI measures is negatively correlated with the degree of fibrosis. In Figure 3A, the correlations between the three different SI based measures included in MPA&aMPA: RE, NLSC_10_ and LMC are shown (Fig. 3A-C). As expected, all measures tended to correlate negatively with increasing 3D-MRE at 33 Hz, with R^2^ = 0.45, 0.50, and 0.71 respectively. Thus, increasing stiffness was correlated with a lower hepatic uptake of Gd-EOB-DTPA. With respect to hepatic fat accumulation (Fig. 3D-E, SI measures, *i.e*., RE, were not correlated with liver fat concentration (R^2^ = 0.008 ^1^H-MRS PDFF; and R^2^ = 0.038 6PD PDFF). With respect to the correlation between RE and plasma ICG (Fig. 3G-H), a trend was observed: ICG-R15 (R^2^ = 0.43), and ICG-PDR (0.16). A negative trend was also observed between 3D-MRE stiffness and liver fat fraction (6PD).

**Figure 3:**
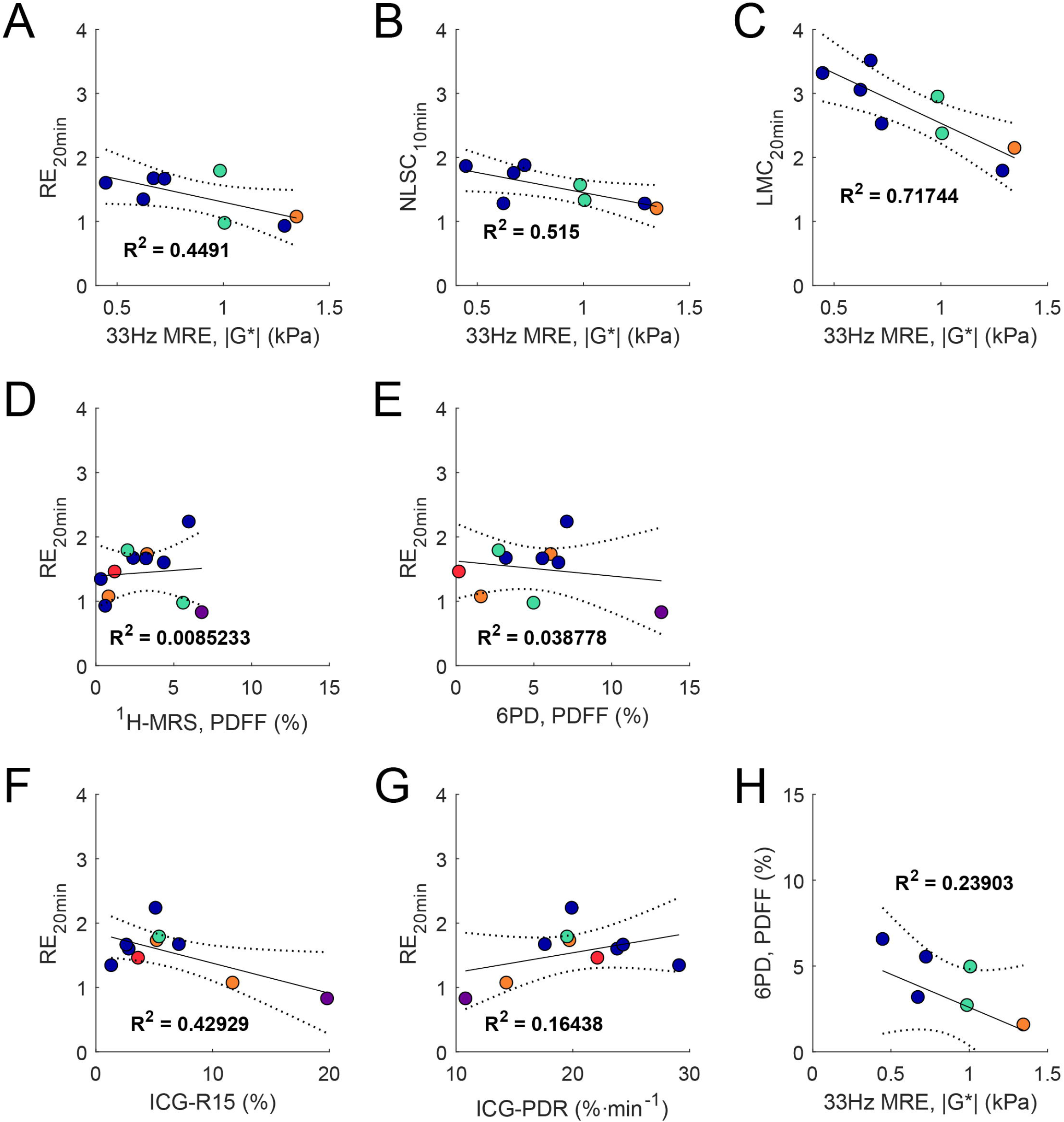
Scatter plots showing SI measures vs. MRE and PDFF. The color indicates the type of resection. **(A) Stiffness vs. Contrast Agent Enhancement**. Scatter plot showing negative correlation relative enhancement (RE) with respect to 3D-MRE stiffness. **(B) Stiffness vs. LSC**. Scatter plot showing negative correlation normalized liver-to-spleen ratio (LSC) at 10 minutes with the 3D-MRE stiffness measurements. **(C) Stiffness vs. LMC**. Scatter plot showing negative correlation liver-to-muscle (LMC) with the 3D-MRE stiffness measurements. **(D) PDFF vs RE**. Scatter plot showing that there is no correlation between PDFF, measured via ^1^H-MRS, and the contrast measurement RE. **(E) PDFF vs RE**. Scatter plot showing that there is no correlation between PDFF, measured via 6PD, and the contrast measurement RE. **(F) ICG-R15 vs RE**. Scatter plot showing negative correlation ICG-R15 with the contrast measurement RE.**(G) ICG-PDR vs RE**. Scatter plot showing positive correlation ICG-PDR with the contrast measurement RE.**(H) PDFF vs. stiffness**. Scatter plot showing negative correlation PDFF with the 3D-MRE stiffness measurements.

### Regional Assessment of Function Using SI-Based Measurements

The Gd-EOB-DTPA based images allowed for regional functional evaluation. Based on ROI placed in each *Couinaud*-segment, a value for all SI based measures could be calculated for each segment (segment 4a and 4b, where combined), assuming that the measures were representative of the function in each segment. In Fig. 4 the RE values from each segmental ROI are shown for all patients. The different groups of resections are shown in different colors, the grey bars show the post-op MRI data. The RE value differed between the different segments for the same patient.

**Figure 4:**
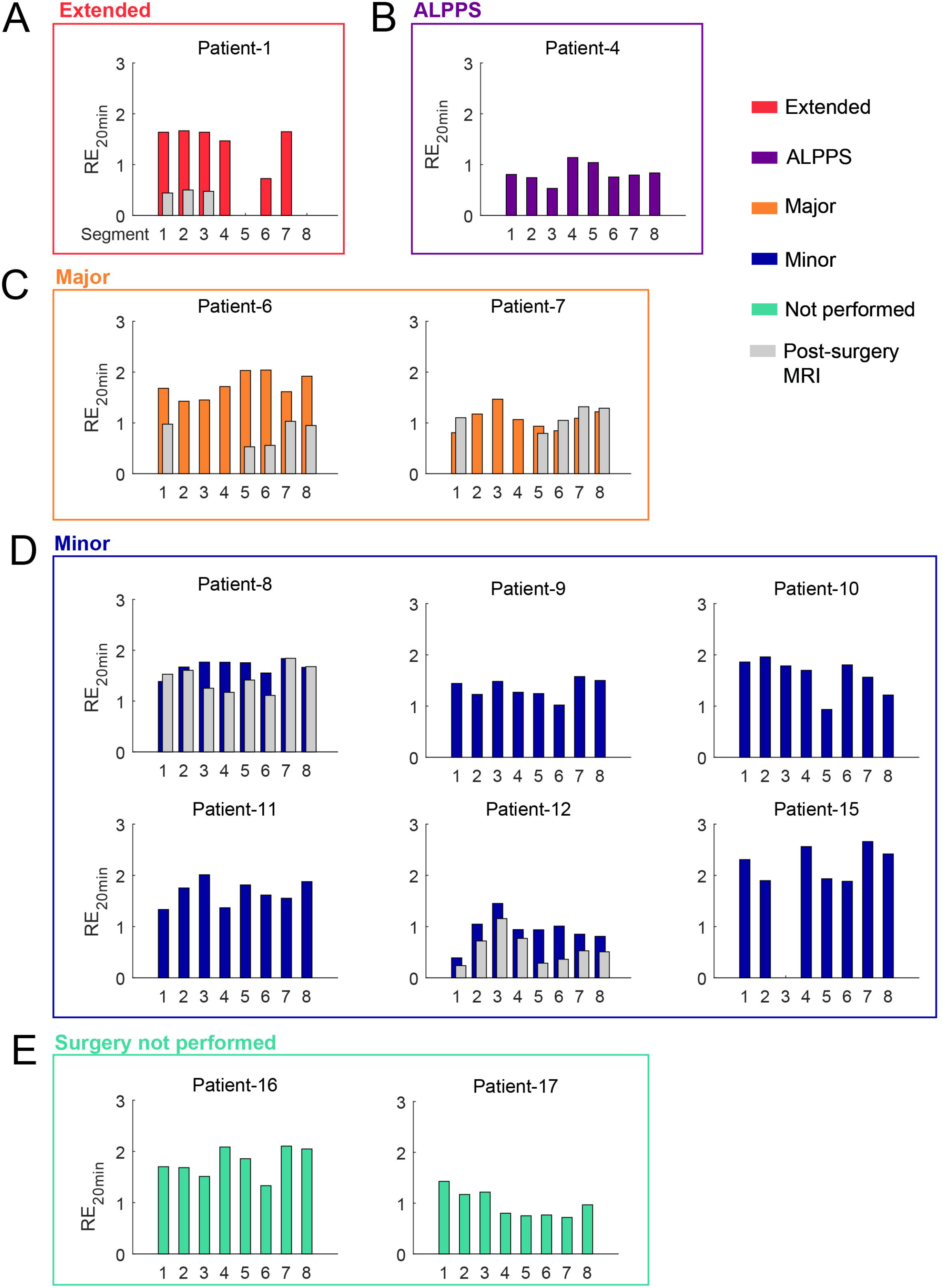
Values of the SI measurement relative enhancement (RE) for ROIs placed in each Couinaud segment for all individual patients. The chart shows the individual values for each segment in pre-op MRI for all patients, in colored bars. White bars represent the RE values in post-op MRI, in front of the pre-op bars. The dotted red line indicated the threshold for the presence of fibrosis (RE = 0.9) presented by Verloh et al. [48]. **(A)** For patients undergoing extended resection; Patient 1. The grey bars are the values for each segment measured during the post-surgery MRI examination. **(B)** For patients undergoing APPLS resection; Patient 4. **(C)** For patients undergoing Major resection; Patient 6, and Patient 7. **(D)** For patients undergoing Minor resection; Patients 8,9,10,11,12 and 15. **(E)** For patients where the resection was not performed; Patient 16 and Patient 17.

For **Patient 1** (Fig. 4A) data were available both for the pre- and post-op MRI examinations. Segments 5 and 8 were excluded as they were occupied by tumor tissue. Pre-op MRI indicates that segment 6 was already below the cut-off for presence of fibrosis (indicating loss in function), of RE = 0.9 presented by Verloh *et al*. [48], while no differences were observed for the other segments. Post-MRI showed that all segments except 1,2 and 3 were resected, and they were all below the cut-off (RE = 0.9, presence of fibrosis). The remaining tissue was very small, and it contributed therefore only little to the global hepatic function. Moreover, Patient 1 later developed PHLF.

**Patients 4, 12 and 17**. The subsequent patients had several segments that fell below the cut-off for presence of fibrosis (RE = 0.9) already during the pre-op MRI; patient 4, 12 and 17 (Fig. 4B, D and E). As previously discussed, Patient 4 had suspected underlaying PC, Patient 12 had confirmed extensive cirrhosis, and Patient 17 had confirmed PC and was thus not resected. We believe that these underlying conditions explain the poor uptake in some of the *Couinaud* segments.

Overall, the post-MRI examination showed a reduction in RE in all segments for all patients, except segment 7 and 8 in Patient 7 and Patient 8 (Fig. C). The largest reduction for a single segment was observed in Patient 6 for segment 5, which had a 74% reduction in RE value. Patient 1 had the largest mean reduction of RE (71%).

## Discussion

We have explored the possible value of using multi-parametric MR as a future clinical decision support tool to enhance the assessment of liver function and status prior to hepatectomy. For this purpose, we recruited a total of 13 patients, who all underwent the complete LIFE protocol, or selected parts thereof, which included both pre-and post-surgery MR-examinations, as well as independent and complementary liver function assessment (ICG-R15, PDR). The very extensive examination protocol encompassed a total of 18 relevant parameters. This work was intended as an exploratory investigation, with an ambition to (i) device a suitable abbreviated MR-protocol, that at the same time would serve as (ii) a tool in pre-operative planning of resective surgery. The selected parameters contribute each to a certain aspect for understanding the complex pattern of liver function and the parenchymal status, which were combined into a single score. In the process to better understand the underlaying cause for patient MPA, and aMPA values, we have provided a detailed overview for each individual patient scenario, for patients undergoing resections ranging from extensive to minor. This work has motivated and encouraged us further to propose an abbreviated MR-protocol.

### Multi-Parametric and Multi-Modality Studies

To our knowledge LIFE is the first study to include such a comprehensive MR-protocol in relation to resective liver surgery, a protocol which included characterization of Gd-EOB-DTPA pharmacokinetics, quantitative MRI, 3D MR-elastography and also conventional function evaluation of liver parenchyma.

In addition to LIFE, there are only a handful of other studies that to some extent explore the use of selected aspects of multi-modality based, or multi-parametric, MRI-protocols in the context of pre-operative assessment. In one study by Wang et al. [53], liver function was investigated using a gadoxetate based protocol in ten colorectal patients undergoing hepatectomy, at several time-points, both before and after surgery. Their protocol included: Gd-EOB-DTPA based DCE-MRI (SI measures included were LMC, LSC, and HUI), but also in addition hepatobiliary scintigraphy (^99m^Tc mebrofenin uptake), as well as ICG function test (R15). However, they did not include MRE, PDFF or other functional measurements. They attribute some promise for DCE-MRI to substitute scintigraphy. Future studies will hopefully provide additional information on the benefits of MR in the pre-operative assessment pipeline. In a planned observational cohort study by Elsharif *et al*. [54], the conventional evaluation with an extended MRI protocol, will be compared with ICG-R15 testing. This may yield valuable knowledge about the benefit of introducing multimodal MR-examinations into the clinical pre-operative-assessment.

### An abbreviated MR-protocol for pre-operative risk assessment?

In this study we have measured an extensive number of MR-derived imaging biomarkers that one could argue in combination would be useful for the characterization of a range of different aspects that affect the intrinsic function of liver tissue in relation to severe liver disease and surgery. However, by necessity such a protocol is relatively time-consuming, and an abbreviated version is therefore naturally desired for applications within a clinical workflow. The use of an abbreviated protocol could increase accessibility and by the same notion also reduce the time these high-risk patients need to spend in the MR-scanner.

Which is then the motivation behind, and approach for, the specific selections of acquisitions in an abbreviated protocol, as is suggested here? Information about the presence and extent of fibrosis could be assessed using a single MRI-parameter, either hepatobiliary specific contrast agent which is an indirect indicator of fibrosis, or direct measurements of tissue biomechanics as probed using 3D MRE. Measurements derived from Gd-EOB-DTPA, *e.g*. RE, have been previously shown to an extent correlate with fibrosis [55], an observation which was also confirmed here. An argument could thus be made that MR-elastography would not be needed in an abbreviated protocol. But on the other hand, one could also argue for the singular use of only using MR-elastography, not only due to the large expense of the hepatobiliary specific contrast agent (which requires up to 30 minutes for completion) compared to the inexpensive and rapid 3D MRE-method (in the order of a minute). The reduction of Gd-EOB-DTPA uptake is mainly due to presence of fibrotic tissue, or perfusion changes caused thereof, that is an indirect measure of fibrosis. Moreover, MR-elastography is also a less invasive alternative, compared with contrast-agent based techniques which involves the use of a xenobiotic agent with potential long term side effects, such as Gd^3+^ accumulation for example in subcortical nuclei of the brain.

In same manner, similar arguments could be made for ICG based functional tests, that correlates to the level of Gd-EOB-DTPA uptake, rapid and inexpensive. However, the latter lacks the possibility for assessing function evaluation on the segmental level, which is deemed to be of major importance for surgical planning. A problem associated with MRE is a potentially large accumulation of iron in the liver, which may occur in some patients, which in turn results in a major signal loss, in particular of significance for GRE-based MRE, at 3 T.

Other measures of importance for determining the functional status of the different segments are liver fat (determined using either MRS or Dixon-techniques), and iron concentration (determined using either R2 or R2*-measurements) [42]. Moreover, energy and anabolic metabolism has been probed in relation to liver disease, suggesting that P-31 Magnetic Resonance Spectroscopy may be a useful, technique for the study of such aspects of the disease development [56].

### Alternative and Expanded Protocols

There are number of alternative imaging techniques such as: scintigraphy, shear-wave ultrasound elastography [57], that have been investigated in the context of hepatectomy. Scintigraphy has shown promise of evaluation of the *function liver remanent* (FLR) [58, 59]. valuation can be performed using e.g., [^99m^Tc] Tc-mebrofenin CT-scintigraphy/SPECT-CT [60]. In addition, scintigraphy has shown some promise in predicting the incidence of PHLF [59, 61]. Additional imaging modalities capable of producing useful information regarding functional aspects might prove to be beneficial, although adding those would contradict the need for abbreviated protocols.

A formal alternative to MRE is ultrasound based elastography, such as transient elastography (TE), *e.g*., FibroScan (Echosense, Paris, France) which offers an assessment of liver tissue stiffness. In HCC patients FibroScan derived liver stiffness has shown potential as marker for predicting PHLF [57, 62]. Moreover, 2D-shear wave elastography (SWE) has recently shown promise as predictor for detecting fibrosis and severe post-hepatectomy complications [63, 64]. Nevertheless, as conventional MRE does have higher diagnostic performance and wider applicability [65], compared to ultrasound based methods and virtual MRE, conventional MRE should primarily be used, if the modality is available.

### Limitations

The presented study has some limitations. One was the data availability for all recruited patients. For example, it was only possible to collect MRE-images for half of the subjects, a condition that was mainly a result of these very ill patients opting out of the examinations, either before inclusion, or into the examination protocol. A more complete set of MRE measurements would allow for a more complete MPA profile for the cohort. Nevertheless, the lack of MRE data did not affect the main scope of our work, as the aim of the work was to focus on individual patients, rather than a group study.

In future work, we will reconsider measurements for liver function, in more detail on a Couinaud-segmental level as it seems reasonable to believe that the function of different segments, or vessel territories, are affected to different extents by the tumor and disease. Although global measures of hepatic function, *e.g*., ICG measurements, in general could also be aiding surgical planning, the segmental level is important in order to deal with the effects to resective intervention. In the future it would be advisable to base all cut-off values on the risk of developing PHLF, or presence of cirrhosis.

## Conclusion

In this pre- and post-observational case-based study ranging from very extensive liver surgery to minor, or none, we have described how a multi-modal MRI examination before hepatectomy could yield useful information for the pre-operative assessment, with a particular focus on a segmental level in individual patients. A multi-modal approach allows for the characterization of several aspects of the remnant tissue. However, more work will be needed to validate the effectiveness and benefit of each parameter, and how to further optimize an abbreviated MR-protocol.

## Data Availability

All data produced in the present study are available upon reasonable request to the authors

## Abbreviations

BCP: Body Composition Profile
HUI: Hepatocellular Uptake Index
MOF: Multiple Organ Failure
MRCP: Magnetic Resonance Cholangiopancreatography
MRE: Magnetic Resonance Elastography
MRS: Magnetic Resonance Spectroscopy
PC: Peritoneal Carcinoma
PD: Point Dixon
PDFF: Proton Density Fat Fraction
PRESS: Point-RESolved Spectroscopy
qMRI: Quantitative MRI
SI: Signal Intensity

